# Causes and consequences of major depressive disorder: An encompassing Mendelian randomization study

**DOI:** 10.1101/2024.05.21.24307678

**Authors:** Joëlle A. Pasman, Jacob Bergstedt, Arvid Harder, Tong Gong, Ying Xiong, Sara Hägg, Fang Fang, Jorien L. Treur, Karmel W. Choi, Patrick F. Sullivan, Yi Lu

## Abstract

**Background:** Major depressive disorder (MDD) is a prevalent and debilitating disorder that has been associated with a range of risk factors and outcomes. Causal pathways between MDD and other traits can be studied using genetic variants as instrumental variables.

**Methods:** A literature review was conducted to identify 201 MDD-associated traits. For 115 traits, there were well-powered genome-wide association study (GWAS) results available that could be used to assess the genetic correlation with MDD. Of these, there were 89 meeting criteria for investigating causal associations in both directions using two-sample Mendelian randomization (TSMR). Of the traits that were not captured by GWAS, 43 could be included as outcomes of MDD using one-sample MR (OSMR). A range of methods and sensitivity tests was applied to gauge robustness of results, together with statistical power analyses to aid interpretation.

**Outcomes:** Moderate to strong genetic overlap was found between MDD and most traits. Support for causal effects of MDD liability were found for circadian, cognitive, diet, medical disease, endocrine, functional, inflammatory, metabolic, mortality, physical activity, reproduction, risk behavior, social, socioeconomic, and suicide outcomes. Most associations were bidirectional, although there was less evidence for diet, disease, and endocrine traits causing MDD risk. Results were robust across sensitivity analyses.

**Interpretation:** This study provides a systematic overview of traits putatively causally related to MDD, confirming previous findings as well as identifying new associations. Our results highlight the importance of MDD as a risk factor cross-cutting across medical, functional, and psychosocial domains and emphasize the need for concerted efforts at reducing this highly prevalent disorder.

## Introduction

Major Depressive Disorder (MDD) is a highly prevalent and debilitating mental disorder. It is crucial to increase understanding in what causes MDD, but also how MDD impacts other aspects of health and well-being. Although treatment provision has increased, prevalence is not decreasing^1^: one in every three women and one in five men experience MDD at some point in their life (Our World in Data^2^). MDD significantly impacts quality of life^3^, accounting for 4.3% of Years Lived with Disability (YLD^4^). It is related to high treatment use, lower socioeconomic outcomes, and adverse clinical outcomes^5^. Also, it is highly co-morbid with a range of medical diseases, most notably with cardiovascular disease^6^ and all disease-related mortality^7^. Moreover, MDD is in the top 3 predictors of suicide ideation (after previous suicide ideation and hopelessness), which in turn is a strong predictor of suicide death^8^. Still, for many outcomes, it is unclear if MDD is causal. For instance, the association with suicide is at least in part driven by psychiatric comorbidity^9^. For other outcomes, there may be reverse causation, with cardiovascular disease for instance being identified as both a risk factor for and outcome in MDD^10^.

Many risk factors have been identified for MDD. For instance, low income, low educational attainment, not having a romantic partner, and loneliness are important social risk factors^11,12^. Psychiatric and medical diseases may also predispose for MDD^5,13^. Diet, substance use, and metabolic traits such as BMI are associated with MDD, too^14^. Childhood trauma and stressful events may be the most robust risk factors for MDD^15,16^. Many risk factors may be subject to bidirectional associations, and there may be other, non-causal pathways driving the association. For instance, the well-established effect of stressful life events might be largely non-causal^17^. It is notoriously difficult to infer causality from observational data, requiring longitudinal or within-family designs.

In contrast, genetic risk factors fixed at conception cannot be affected by reverse causation. The heritability of MDD has been estimated at 30-50% in family-based studies^5,18^. Genome-wide association studies (GWAS) have been able to identify numerous common variants that contribute to the disorder^19,20^, explaining up to 7% of the variance^21^. Using genetic variants associated with a trait as instruments to capture it alleviates some of the difficulty in testing causal, directional associations. Such Mendelian Randomization (MR) analysis can support causal interpretations if instrument variants have an effect on the outcome through their association with the risk factor, without having a relationship with the outcome or unmeasured confounders. Previous broad-ranging MR studies in MDD have largely focused on modifiable risk factors for MDD, or the causal link between MDD and disease outcomes. Support has been found for causal effects in both directions between MDD and health behaviors, cognitive measures, social behaviors, cardiovascular disease, and metabolic traits^6,22–25^. In this study, we include traits that have been associated with MDD in previous observational studies, for which large-scale GWAS data was available. In addition, we assess causal effects of MDD on outcomes for which there are currently no GWAS data available (including outcomes related to daily functioning), relying on one-sample MR techniques. This approach allows us to give a systematic overview of putatively causal associations and will inform which traits should be targeted in prevention and intervention of MDD and its consequences.

## Methods

We used instruments from GWAS summary statistics to assess putative causal effects between MDD and a broad selection of traits associated to MDD in observational studies. For MDD, we used the newest publicly available GWAS that identified 243 risk loci and explained 7% of the variability in MDD^21^. We used summary statistics leaving out the UK-Biobank to limit sample overlap, with 166,773 cases and 507,679 controls. We investigated genetic correlations and one- and two-sample MR associations with a wide range of literature-identified traits.

### Trait selection

The selection of risk factors and outcomes of MDD was guided by literature search and restricted by the availability of GWAS with sufficient sample size and power. To identify risk and outcome factors we relied on a selection of review papers^11–15,26–29^. For trait groups suggested by the review papers (e.g. gastrointestinal disease) we searched the literature additionally for specific reports of associations (e.g., gastroesophageal reflux disease). The considered traits (164 traits), their literature source, as well as any available corresponding GWAS sources are summarized in Table S1. Subsequently, we searched corresponding GWAS sources for each trait, and included them if they met the following criteria: N≥10,000 for continuous traits, or N cases≥5,000 for binary traits; 3% SNP-based heritability; and at least 5 GW-SNPS. Though not formally pre-registered, these criteria and all downstream methods were established before conducting any analyses. Criteria are explained in more depth in Supplementary Information 1.

### LD Score regression

SNP-based heritability (h^2^), which was used as a selection criterion for the GWAS traits, was derived using LD Score Regression^30^. We used the sample prevalence as an approximation for the population prevalence for the computation of h^2^ on the liability scale for binary traits. The use of sample prevalence as an approximation for population prevalence was deemed sufficient because we only used h^2^ as a selection criterion and were not interested in the heritability per se. Subsequently, we used LD Score Regression to estimate the genetic correlation between MDD and each of the other traits. To correct for multiple testing, we applied a conservative Bonferroni correction of *p* = 0.05/ 115 (the number of tested traits), resulting in a threshold of *p* < 4.4E-4. Although causal associations may exist in the absence of genetic overlap, for polygenic traits it is expected to be accompanied by corresponding genetic correlation^31^. To investigate this, we assessed the relationship between the genetic correlation and the MR effect estimate.

### Two-Sample Mendelian Randomization (TSMR)

Next, the putative causal associations between MDD and the traits were tested using Two-Sample MR (TSMR). We assessed the effects in both directions, with MDD as exposure and as outcome. In TSMR, both the exposure and the outcome are proxied by GWAS summary statistics. Instruments were created by selecting the independent (clumping at R^2^ < 0.001, distance < 10,000 kb) genome-wide significant SNPs (*p* < 5E-8) that were present in both the exposure and outcome trait GWAS. When there were fewer than 10 instruments remaining after this procedure, we increased the *p*-value threshold until there were 10 to maximum *p* < 1E-5. After aligning with the outcome data the final number of instruments could still fall below 10, but was never smaller than 5. We used the Two-Sample MR R-package^32^ to conduct Inverse Variance Weighted (IVW) meta-analysis of effects, which is the standard in the literature and is the estimate that is reported in the main text. We use a broad selection of state-of-the-art alternative methods, sensitivity analyses, and robustness checks, including Generalised Summary-data-based MR (GSMR), Latent Heritable Confounder (LHC) MR, and Steiger-filtering (Supplementary Information 2).

Although it is often recommended to avoid causal language when describing results from MR analyses (i.e., preferrable would be to say ‘liability to X causes risk for Y’), we sometimes use causal language for brevity and clarity throughout the paper, following previous recommendations^33^. Although the scale and specific features of this study did not allow for full adherence to the STROBE checklist (a well-established protocol developed to ensure the quality of MR reports^34^), we have followed it as much as possible throughout the analyses (Supplementary Information 3).

### One-Sample Mendelian Randomization (OSMR)

Subsequently, we performed One-Sample Mendelian Randomization (OSMR) in the UK-Biobank with MDD as exposure and electronic health record- and survey-based outcomes. The OSMR analysis is conducted using individual-level data in two stages. First, SNPs are used as instruments to predict the exposure. Then, the genetically predicted exposure variable is used to predict the outcome. The advantage with respect to TSMR is that traits that have not been captured by GWAS can still be used as outcome. The advantage compared to standard polygenic risk score analysis is that assumptions are tested to assess if results are in line with a causal interpretation. We focus on the traits that the literature review suggested were of importance, but could not be tested with TSMR due to the lack of suitable GWAS sources. The selected outcomes and the MDD exposure are described in Supplementary Information 4.1.

For the OSMR analyses we used a set of two-stage methods as well as MR methods that are also used in the two-sample context. To create the MDD instrument, we created a polygenic risk score (PRS) with Plink^35^ based on the MDD GWAS leaving out UK-Biobank to prevent bias due to sample overlap. Effects of birth year, sex, and the first 10 principal components capturing genetic ancestry differences were regressed out of the PRS. We used the same SNP selection criteria as for TSMR and created per-individual weighted sum scores of these SNPs. Because this score included only a small subset of SNPs and would have limited power, we repeated the analyses with a PRS created with LDpred2^36^ (Supplementary Information 4.2). For each trait, we then computed the OSMR causal estimate using two-stage least squares (2SLS) regression as implemented in the ivreg R-package, relying on medical records of MDD diagnosis as the observed variable (N=28,861 cases, N=103,065 controls, average birth year M=1950, SD=8.1) and the PRSs as instrumental variables. Wu-Hausman diagnostics were used to test endogeneity: if all regressors are exogenous, the 2SLS and OLS estimates are consistent. Thus, a significant Wu-Hausman indicates that one or more regressors is endogenous and the 2SLS outcome is preferrable over the OLS estimate. Instrument strength F was derived in a parallel manner as for TSMR (Supplementary Information 4.3). As a comparison, we also derived the ordinary least squares (OLS) regression estimates for the effect of observed MDD diagnosis on the outcomes (Supplementary Information 4.4). To be able to conduct sensitivity analyses including IVW, weighted median, and MR Egger, we also computed the relationship between each instrument SNP and the outcomes in UK-Biobank using Plink association analysis (Supplementary Information 4.5).

### Power and multiple testing correction

We used power calculation to prioritize traits which were more likely to yield reliable results in the TSMR and OSMR^37^. For TSMR, we used the R-app https://sb452.shinyapps.io/power/^38^. For OSMR, we used the app https://shiny.cnsgenomics.com/mRnd/ ^39^. To arrive at an approximation of the range of effect sizes in observational studies, we used effect sizes as reported in the literature that we used for trait selection. We chose a plausible effect size range of β=0.1-0.3/ OR=1.1-1.3 (based on literature summarized in Table S1). We used our own observations of the variance explained by the instruments (R^2^) in the exposure as input for the power analyses.

We used false discovery rate (Benjamini-Hochberg FDR) *p*-values to correct for the number of traits tested, separately for the OSMR and TSMR analyses. For interpretation of the TSMR results, we weighed the consistency of effect sizes across different sensitivity analyses rather than only relying on any single *p*-value falling below the *p_FDR_ =* .05 threshold.

## Results

### Trait selection

In the literature review, 201 relevant traits associated with MDD were identified (Fig. 1, Table S1). In many cases, the associations were framed as bidirectional, such that MDD could plausibly be the exposure as well as the outcome for the trait of interest. Of the identified traits, 133 were captured in a publicly accessible GWAS (Table S2). The genetic correlation with MDD could be assessed for 115 traits. From those, 25 were excluded for TSMR, because they had insufficient SNP-heritability or had too few genome-wide significant SNPs (Fig.1, Table S2. We excepted traits related to suicide which were kept because they are pertinent outcomes of MDD (though results should be interpreted cautiously). In the end, 89 traits were included for TSMR. Of the traits that did not meet the criteria for TSMR, 43 were captured (in a total of 48 variables) in the UK-Biobank data and could be included in the OSMR analysis (Table S3).

**Fig 1.**
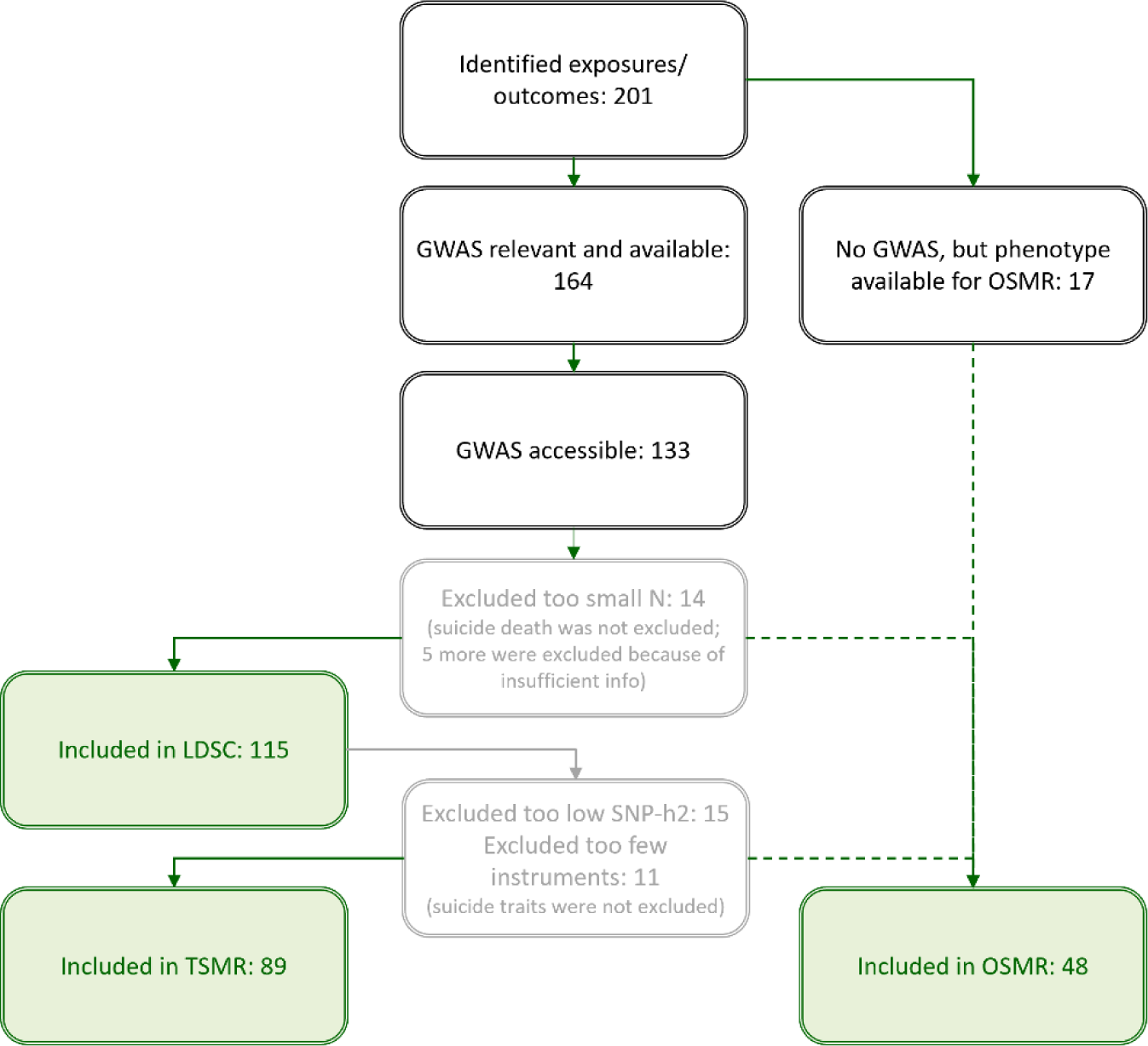
Trait selection procedure with the final number of traits included in each analysis.

### Genetic correlations

In Fig. 2 the genetic correlations between MDD and the selected outcome traits are displayed. The strongest associations (in positive or negative direction) were found for subjective health and pain-related disease, trauma and stress, divorce, loneliness, suicide, and functional outcomes. Most other traits showed moderate (.20-.50) levels of genetic overlap. Detailed results are in Table S4.

**Fig 2.**
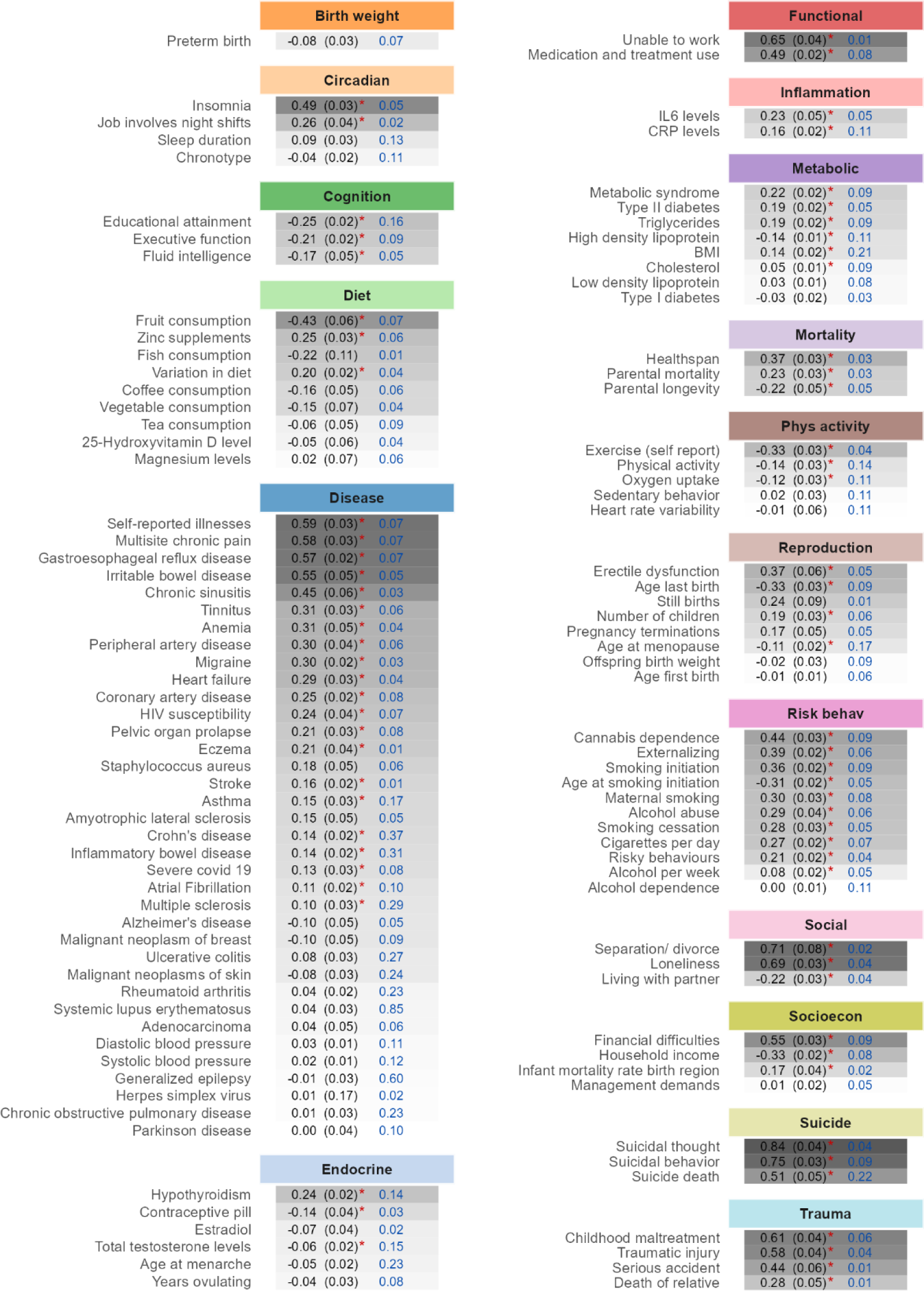
Genetic correlation (standard error) between MDD and manually categorized traits. Asterisks indicate a significant correlation at a Bonferroni-corrected *p* < 4.4E-4 (for 115 traits). In blue the SNP-based heritability is reported as computed using LDSC (on the liability scale for binary traits, using sample prevalence to stand in for population prevalence). The shading indicates the strength of the correlation, with the strongest correlations reported on top and in the darkest shade.

### Two-Sample Mendelian Randomization (TSMR)

The main results (IVW estimates) from the TSMR are summarized in Fig. 3, along with the estimated explained variance by the instrument in the exposure R^2^ and the statistical power. Among the 89 disorders and traits studied, we found more support for MDD having a causal effect on a trait (57 traits; average absolute effect size M=0.18, average SE=0.05) than for traits having causal effects on MDD (24 traits, M=0.13, SE=0.06). Of the 24 traits with support for an effect on MDD there was only one that did not also show an effect in the other direction. There was a sizable association between the genetic correlation and the effect size from TSMR, for the analyses with MDD as exposure *r*=0.81 (95% confidence interval [CI]= 0.73-0.87) and with MDD as outcome *r=*0.59 (CI=0.43-0.71; Fig. S1). Average power was 57-87% for the analyses with MDD as exposure and 64-94% with MDD as outcome under hypothesized ‘true’ effect sizes β=0.1-0.3 (or OR=1.1-1.3). A generalized power curve showing which sample sizes and effect sizes would be needed to achieve sufficient power is displayed in Fig. S2. The effect sizes should be interpreted in light of these power estimates, although there was no clear positive relationship between power and the MR effect size (Fig. S3).

**Figure 3.**
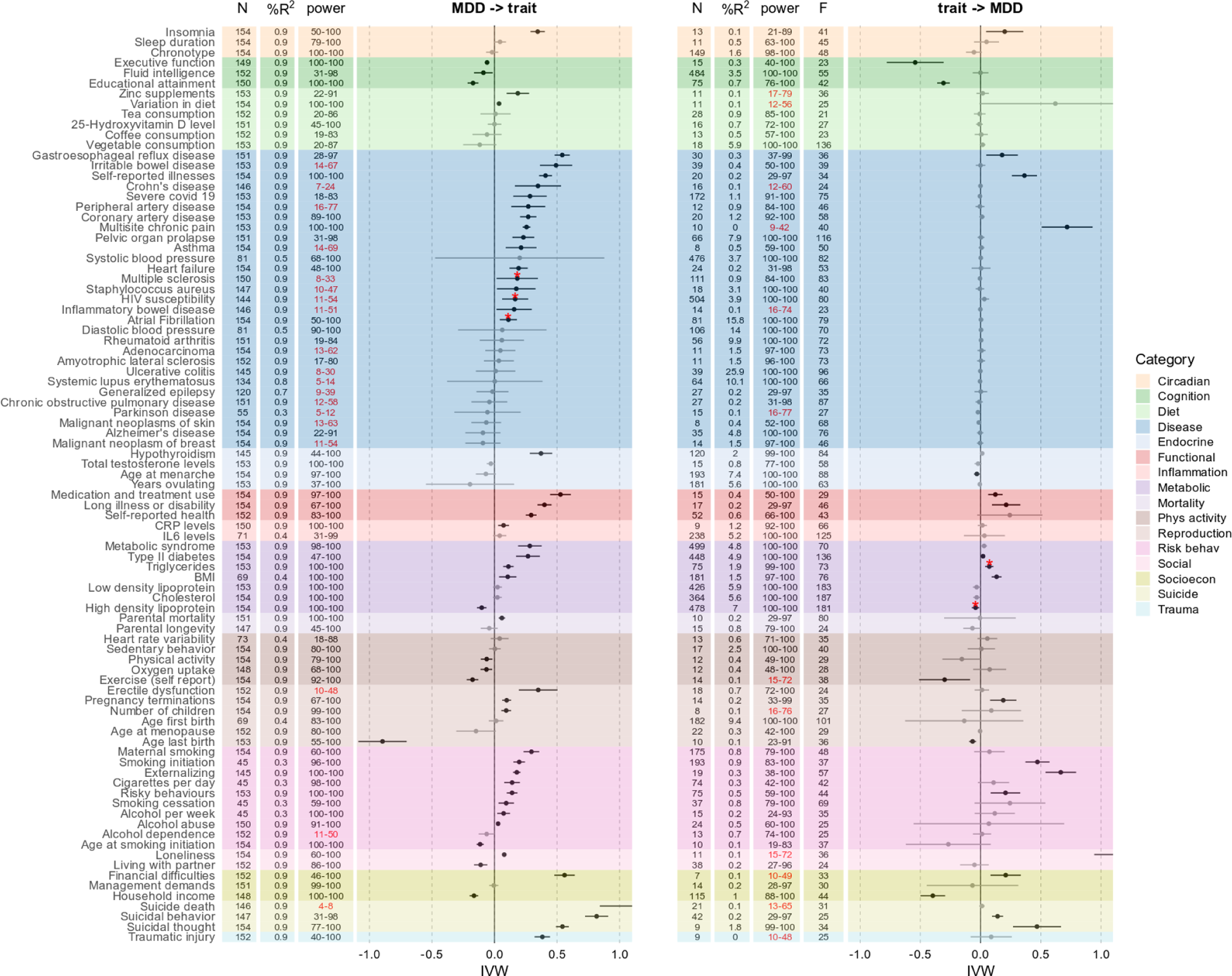
Putatively causal effects of MDD on 89 other disorders and traits (left) and vice versa with MDD as outcome (right). The forest plots show the IVW estimate with 95% CI. Estimates printed in black survive correction for multiple testing (*pfdr* <0.05). Estimates highlighted with an asterisk are significant estimates that showed evidence of horizontal pleiotropy (at a conservative MR Egger intercept *p*<.05). The columns show the number of instrument SNPs for each analysis (“N”), the percentage variance explained by the instruments in the exposure (“%R^2^”), the achieved level of power under an effect size of β=0.1-0.3/ OR=1.1-1.3; “power”; estimates where the upper limit fell short of 80% printed in red), and the instrument strength of the exposure (“F”). In the left panel, the exposure is always MDD, but R^2^ still varies somewhat due to the variation in the number of SNPs present in the outcome summary statistics (F varied little [F=40.1-42.6] and is not shown). Note that the estimate of the effect of loneliness on MDD was out of the range of the plot (IVW=2.17, SE=0.63). Also note that the summary statistics for anemia, atrial fibrillation, chronic sinusitis, severe covid-19, cholesterol, alcohol dependence and cannabis dependence were based on analyses in multi-ancestry populations (with the largest sub group being European).

For the *circadian* traits, we found support for a positive bidirectional effect of MDD on insomnia, such that genetic liability to MDD increased insomnia risk, and vice versa, while there was limited evidence for a similar effect on sleep duration or chronotype from either direction. Likewise, in the *cognition* category, there was support for a bidirectional causal negative effect for executive function and educational attainment, whereas there was a negative effect of MDD on fluid intelligence which was not observed in the other direction. For *diet*, the statistical power was mostly limited, but we still detected small positive effects of MDD on taking zinc supplements and dietary variation. There was evidence for a causal effect of MDD on a number of *diseases*, including gastrointestinal diseases, COVID-19, chronic pain, a number of cardiovascular diseases, and others (the effects on multiple sclerosis, HIV susceptibility and atrial fibrillation were pleiotropic and should not be interpreted). There was little evidence for an effect on cancer or neurological disease, although power was limited for many of these analyses. In the other direction, there was only support for a causal effect of chronic pain, the self-reported total number of somatic illnesses, and gastroesophageal reflux disease. For *endocrine* traits, MDD increased risk for hypothyroidism, and a younger age of menarche marginally increased MDD risk. There were bidirectional causal associations with most *functional* outcomes. For the *inflammatory* traits, there was only evidence for a causal effect of MDD liability on elevated CRP levels. In the *metabolic* category, there was evidence that MDD increased the risk for all outcomes except low-density lipoprotein and total cholesterol. In the other direction, there was evidence for an effect of BMI and a small effect of type II diabetes. MDD liability had a positive effect on parental *mortality*. In the *physical activity* category, there was a negative effect of MDD on physical activity, exercise and oxygen uptake (a measure of fitness), and a negative effect of exercise on MDD. In the *reproduction* category, there was evidence for a positive effect of MDD on erectile dysfunction and the number of children, and bidirectional effects with pregnancy terminations and a younger age at last giving birth. There was support for a causal effect of MDD on all *risk behavior* outcomes except alcohol dependence (this analysis was underpowered). In the other direction, there was evidence for a causal effect of externalizing behavior, smoking initiation, and risky behaviors on MDD. For the 2 *social* traits, MDD lowered chances of living together with a partner and increased loneliness, while loneliness had an out-of-bounds effect on MDD (IVW=2.17, SE=0.63, *p_FDR_*=.003). In the *socioeconomic* category, there was evidence for a bidirectional association with a lower income and a higher chance of experiencing financial difficulties. There were effects of MDD liability on all *suicide* outcomes with an out-of-bounds effect on suicide death (IVW=1.12, SE=0.14, *p_FDR_*=.2E-15, though power was 4-8%), and smaller but significant effects of suicidal behavior and thought on MDD. Finally, MDD increased the chances of self-reported *traumatic* injury, whereas traumatic injury had no effect on MDD.

The full TSMR results including all sensitivity analyses are presented in Table S5. Steiger-filtered IVW, MR Egger, simple mode, weighted median and mode estimates were consistent with the IVW, though sometimes significance was reached in one, and not the other. Results were more pronounced in the GSMR and MR-PRESSO analyses (Fig. S4-S5), but confidence intervals were overlapping with the IVW confidence intervals in all analyses. The LHC analysis, that aims to correct for heritable confounders and sample overlap, mostly showed effects in the same direction, though with less precision. Finally, few trait combinations showed evidence of pleiotropy, as evidenced by the MR Egger intercept (asterisks in Fig. 3). Overall, the sensitivity analyses were in line with a causal interpretation of the IVW.

### One-Sample Mendelian Randomization (OSMR)

We conducted OSMR to test the effect of MDD on 48 outcomes for which no GWAS were available. Compared to TSMR, the statistical power of OSMR was much more limited (average 7-27% assuming effect sizes β=0.1-0.3 or OR=1.1-1.3) because the instrument SNPs explained only minute amounts of variance in the MDD exposure (R^2^=0.08%, F=0.58). Nevertheless, the OSMR diagnostics indicated that the MDD instrument did not suffer from weak instrument bias (for the significant findings at *p_FDR_*<.05, average weak instrument test *F*=39.9, average *p=*3.5E-6).

Despite the overall limited power in the OSMR, we found evidence that MDD has an effect on a range of functional outcomes, increasing pain, problems with daily life, and hospitalizations, while reducing health satisfaction (Table S6). Results are presented in Fig. 4 alongside the ordinary least squares (OLS) regression results relying on observational data only (MDD diagnosis predicting each outcome). The strongest OLS associations were found in the disease and functional outcomes category, with protective effects of MDD on disease outcomes. These protective effects are not in line with previous literature and findings from TSMR. They could be the effect of censoring in the hospital record data or, speculatively, be driven by a lower likelihood of an MDD being diagnosed in the presence of a medical disease that can be an alternative explanation for the observed symptoms. Regardless, the OSMR findings suggest that the protective effects of MDD on disease are not causal, while the negative impact on functional outcomes is. In addition, according to the OSMR results, genetic liability to MDD decreased the likelihood of being a fruit consumer and of having an accident, increased the chance of alcohol-related mortality, living in a socially deprived region, engaging in self-harm, and of experiencing partner violence.

**Figure 4.**
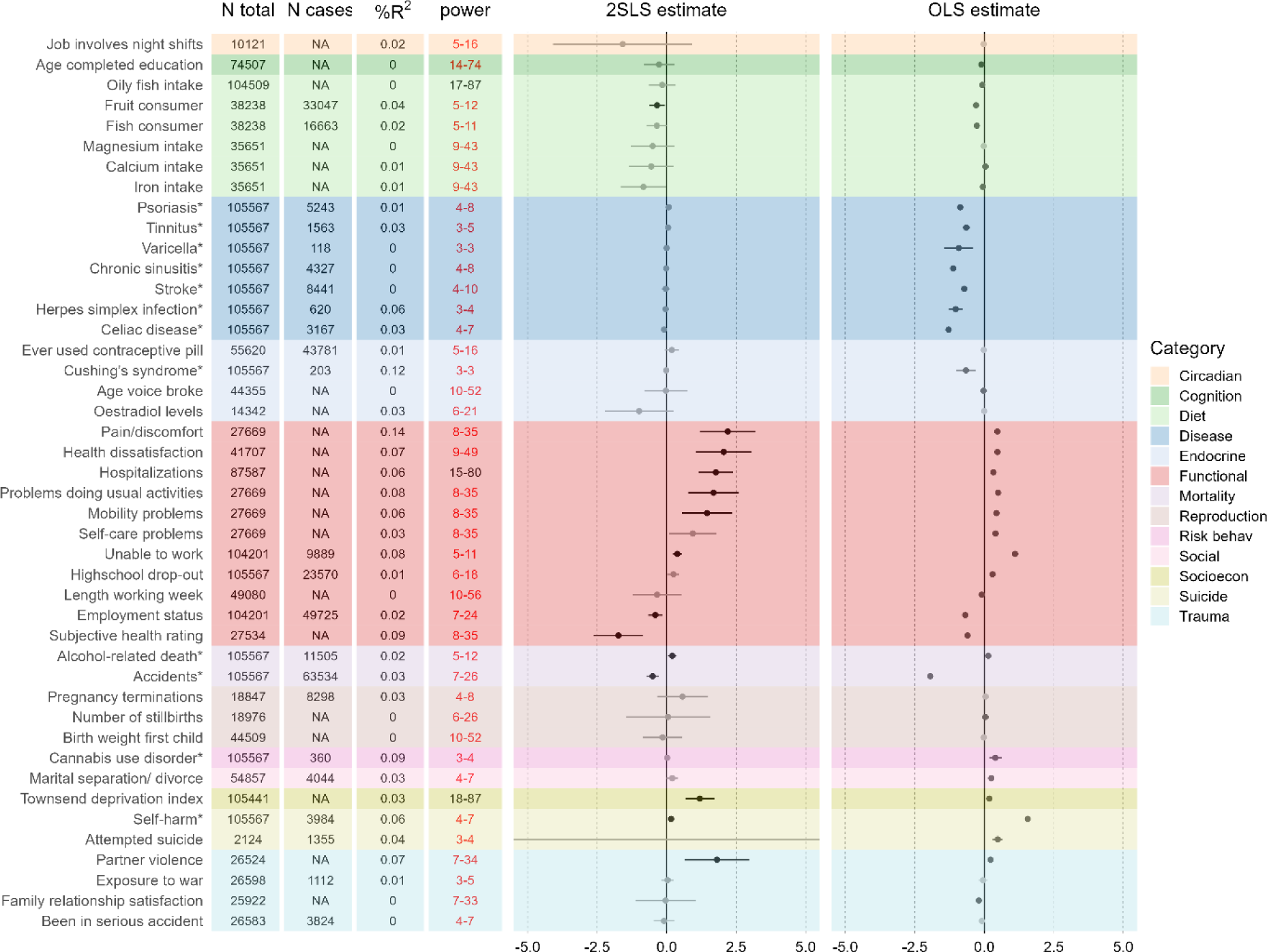
Results from the follow-up OSMR analysis for traits for which there was no GWAS available suited for TSMR. The instrument was the polygenic risk score for MDD, the outcomes were electronic health recorded ICD-codes (marked with *) or survey-based phenotypes from UK-Biobank. Sample size (N), the number of cases for binary phenotypes (N cases; ‘NA’ for continuous phenotypes), explained variance by the instrument in the outcome (R^2^), and the achieved level of power under an effect size of β=0.1-0.3/ OR=1.1-1.3; “power”; estimates where the upper limit fell short of 80% printed in red) are given in the columns. NOTE: R^2^ is used here to represent variance explained in the outcome rather than the exposure (as in the TSMR analyses) because the exposure was the same across analyses in the OSMR. The interpretation is accordingly different, such that it represents instrument strength in the TSMR, and effect size in the OSMR.

Diagnostic tests and sensitivity analyses indicated that, for most analyses, the OSMR results were robust (Table S6). The Wu-Hausman diagnostic indicated that the data were more in line with the 2SLS OSMR estimate (with a causal interpretation) than the OLS (non-causal; for the significant relationships, average Wu-Hausman=20.0, *p*=.004; full OLS results are in Table S7). The sensitivity analyses based on per-instrument association tests were generally in line with 2SLS, without strong evidence for pleiotropy for the outcomes that showed a significant effect in the main OSMR analysis, and generally consistent effects in the IVW and weighted median analysis. The 2SLS analyses using the LDpred2 instrument PRS showed similar results, but the causal estimates had smaller confidence intervals, so that almost all effects reached significance (Fig. S6 and Table S6). LDpred2 results are discussed in more detail in Supplementary Information 5.

## Discussion

Using a literature-based selection of 115 traits captured in GWAS and 48 additional traits for which no GWAS was available, this study assessed the genetic overlap between MDD and its risk factors and outcomes, and applied a range of MR methods to investigate putatively causal associations. We observed genetic overlap between MDD and traits from all categories, with the strongest correlations observed for subjective health and pain-related disease, stress, loneliness, suicide, and functional outcomes. In addition to such pleiotropic associations, there was also support for a causal effect of MDD liability on circadian, cognitive, diet, disease, endocrine, functional, inflammatory, metabolic, mortality, physical activity, reproduction, risk behavior, social, socioeconomic, suicide, and traumatic injury. In the other direction (in the TSMR analyses only), there were fewer associations, with less evidence for diet, disease, and endocrine traits causing MDD risk. The effects were overall robust across sensitivity analyses and consistent in different MR methods, and, to the extent that this can be assessed using current tools, most were robust to violation of the assumptions of MR.

Our TSMR results were more in line with a causal effect of MDD liability on other traits than vice versa, although for 24 traits we found evidence that they act both as consequence and cause to MDD. Especially in the medical disease category, most traits were consequences of MDD, which follows results from an earlier report based on the UK-Biobank^24^, but is contrary to other work suggesting the relationship with medical disease is mostly bidirectional^40^. Potential exceptions to this pattern of unidirectional effects of MDD on disease are liability to chronic multisite pain, number of illnesses, and gastroesophageal reflux disease, that also had a putatively causal effect on MDD. The first two traits were measured through self-report and may be confounded by a third variable influencing self-report as well as MDD, such as negative recall bias, even though this is not suggested by the sensitivity analyses (i.e., no evidence for pleiotropy). Regardless, these findings suggest that there may be something specific about these traits that impacts MDD in a way that other disease traits do not, potentially through psychological and health behavior mechanisms^40^. The OSMR results underline the negative consequences of MDD and show a significant impact on daily functioning. The wide range of adverse effects of MDD illustrates its universal negative impact across domains that could be mediated through chronic stress, inflammation, and unhealthy behavior associated with the condition.

The TSMR analysis shows evidence for many more causal associations than previous studies, which is likely due to the inclusion of a broader range of phenotypes and the use of the newest, high-powered summary statistics for MDD as well as many other traits. For instance, to our knowledge, previous studies have not found consistent evidence for an effect of MDD on diet or infectious disease^22,24,41^. Also, by adding OSMR analyses we were able to show putatively causal effects of MDD liability on traits that have not been (reliably) captured in GWAS. For instance, this study may be the first to use this method to support the interpretation that MDD causally impacts quality of life. OSMR has been little leveraged in previous studies due to the limited power associated with this method^42^. By relying on a strong GWAS source and a broad range of OSMR tools, we show that OSMR can still be used to show relative support for a causal interpretation.

Interpreting the nature of these causal associations is in many cases not straightforward. For instance, depressive mood is a well-established symptom of hypothyroidism, but we find evidence for hypothyroidism causing MDD, and not the other way around (opposing findings from^43^). This could be due to the phenotypes used in the source GWASs; especially in the case of MDD it has been argued that broad, inclusive definitions may dilute the genetic signal and amplify unimportant correlations^44^. Even if methodological and statistical issues are likely part of the explanation (e.g., there could be some residual pleiotropy not picked up in the sensitivity analyses, especially given MDD’s highly polygenic nature), this paper should be viewed as a starting point to explore such potential pathways to further elucidate the etiology of MDD as well as its consequences.

This study has some key strengths and limitations to consider. One important strength is the wide selection of traits, methods, sensitivity analyses, and robustness checks employed. The MR method is continuously being expanded and improved so that a comprehensive, robust, and flexible toolbox has become available. Another key strength is the integration of statistical power, in the selection and creation of instruments, as well as in the interpretation of the findings. Our comprehensive, literature-based selection of traits is a strength as it provided a broad overview of the important risk factors and outcomes. However, it also resulted in limited space to develop hypotheses and interpretations for each tested association, a practice that is recommended in MR research^37^. Likewise, we have not triangulated our findings using different non-MR methods and observational datasets^45^. Our application of stringent selection criteria for trait inclusion ensured statistical power, but has led to the exclusion of key known risk factors for MDD, especially in the trauma category. Even in the light of these stringent criteria, there was a power imbalance in many cases, such that the MDD GWAS was better powered than the GWAS for the other trait, which could partly explain the pattern where we observed more effects of MDD than vice versa. Another limitation lies in the fact that we did not stratify outcome summary statistics for exposures that are measured only in a subset of individuals. For instance, the effect of alcohol dependence should ideally be tested in alcohol-exposed individuals, and the effect of ovulation should only be assessed in women (although, on the other hand, stratification can lead to collider bias)^46^. Relatedly, most binary traits included in this study are dichotomizations of underlying continuous traits. This can lead to violation of the assumption that the instrument can only influence the outcome through the exposure: it can have an effect via the underlying continuous trait, without changing the level of the binary variable^47^. Although there is no real solution to this issue, the interpretation of results relying on dichotomized exposures as pertaining to the liability to this trait is still valid.

While mindful of these caveats, we conclude that this study provides evidence for putatively causal effects of a wide range of traits on MDD liability, and vice versa. We offer support for the causal nature of known risk factors that can provide efficient targets for prevention and intervention efforts, including a wide range of health behaviors, loneliness, cognitive traits, pain, and gastroesophageal reflux disease. Particularly relevant to highlight are potentially modifiable risk factors, including BMI, loneliness, exercise, and (maternal) smoking. The evidence for a causal effect of MDD liability on a plethora of outcomes, including disease, suicide, and mortality, emphasizes the need for better treatment of this prevalent and debilitating disorder. Our results demonstrate the key role of MDD as risk factor cross-cutting across medical and psychosocial domains. As such, this study provides strong weight to the call for concerted action aimed at decreasing this highly prevalent and debilitating disorder^48^.

## Acknowledgments

This work was supported by the US National Institutes of Mental Health (R01MH123724), the European Union’s Horizon 2020 Research and Innovation Programme (CoMorMent project; Grant #847776) and the European Research Council (grant agreement ID 101042183). The OSMR analyses have been conducted using the UK Biobank Resource under Application Number 22224. They were enabled by resources in project sens2017519 provided by the Swedish National Infrastructure for Computing (SNIC) at UPPMAX, partially funded by the Swedish Research Council through grant agreement no. 2018-05973, and in project 2023/5-402 provided by the National Academic Infrastructure for Supercomputing in Sweden (NAISS) at PDC, partially funded by the Swedish Research Council through grant agreement no. 2022-06725. JAP was funded by Forte - The Swedish Research Council For Health, Working Life And Welfare (agreement 2022-00814), the US National Institutes of Mental Health (R01MH123724), and by the Amsterdam University Medical Center Postdoc Career Bridging grant (27527). KWC was supported in part by funding from the National Institute of Mental Health (K08MH127413). PFS gratefully acknowledges support from the Swedish Research Council (Vetenskapsrådet, award D0886501) and the US National Institutes of Mental Health (R01s MH124871, MH121545, and MH123724). YL was supported by the European Research Council (grant agreement ID 101042183).

## Data availability

All GWAS summary statistics used are publicly available. The UK-Biobank data source is accessible to all researchers upon application. All code and scripts used for this project will be shared on https://github.com/KI-PGI.

