## Supplementary figures for "Causes and consequences of major depressive disorder: An encompassing Mendelian randomization study"

### Slide 1
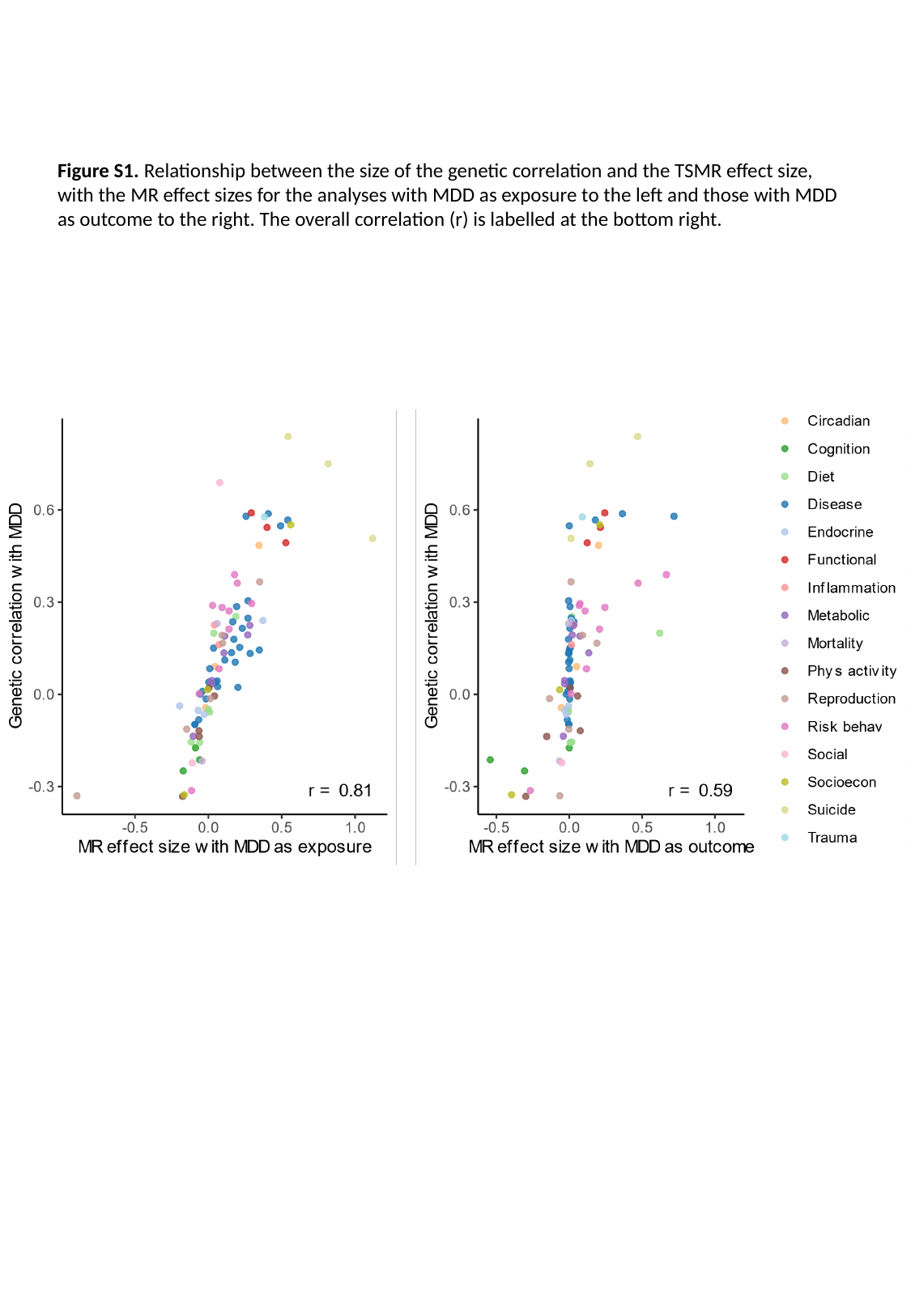

Figure S1. Relationship between the size of the genetic correlation and the TSMR effect size, with the MR effect sizes for the analyses with MDD as exposure to the left and those with MDD as outcome to the right. The overall correlation (r) is labelled at the bottom right.

### Slide 2
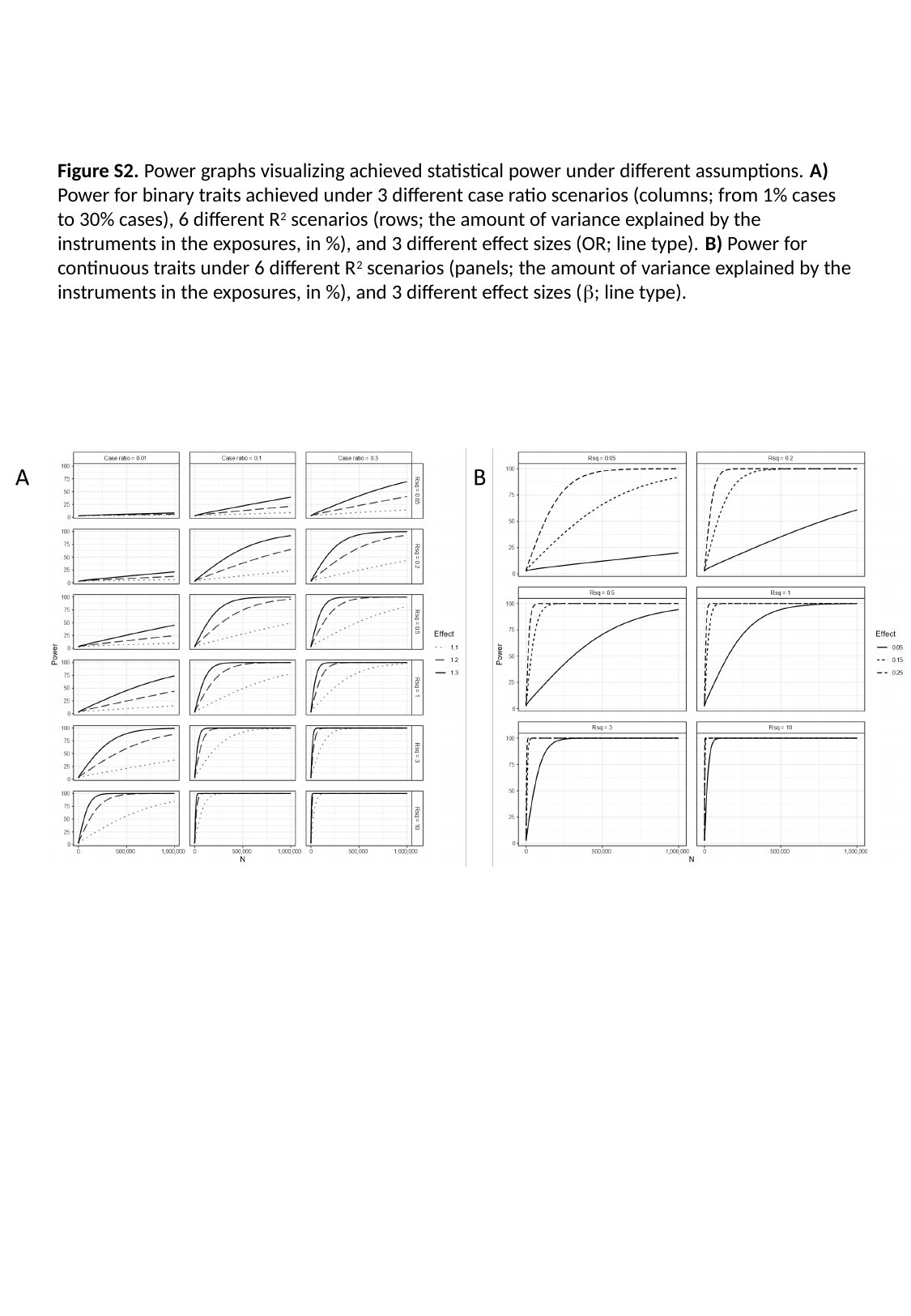

Figure S2. Power graphs visualizing achieved statistical power under different assumptions. A) Power for binary traits achieved under 3 different case ratio scenarios (columns; from 1% cases to 30% cases), 6 different R^2^ scenarios (rows; the amount of variance explained by the instruments in the exposures, in %), and 3 different effect sizes (OR; line type). B) Power for continuous traits under 6 different R^2^ scenarios (panels; the amount of variance explained by the instruments in the exposures, in %), and 3 different effect sizes (β; line type).

### Slide 3
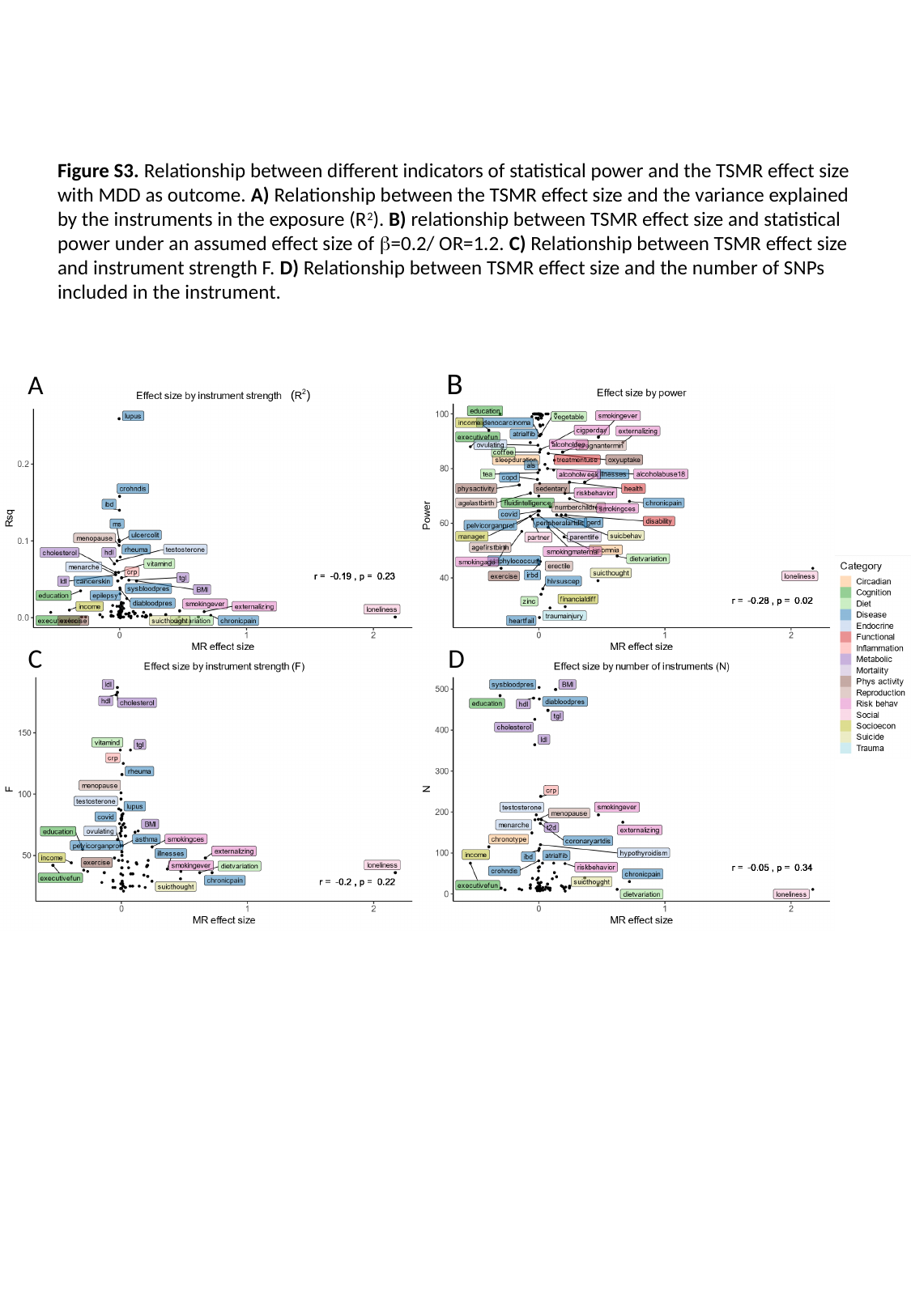

Figure S3. Relationship between different indicators of statistical power and the TSMR effect size with MDD as outcome. A) Relationship between the TSMR effect size and the variance explained by the instruments in the exposure (R^2^). B) relationship between TSMR effect size and statistical power under an assumed effect size of β=0.2/ OR=1.2. C) Relationship between TSMR effect size and instrument strength F. D) Relationship between TSMR effect size and the number of SNPs included in the instrument.
B
A
C
D

### Slide 4
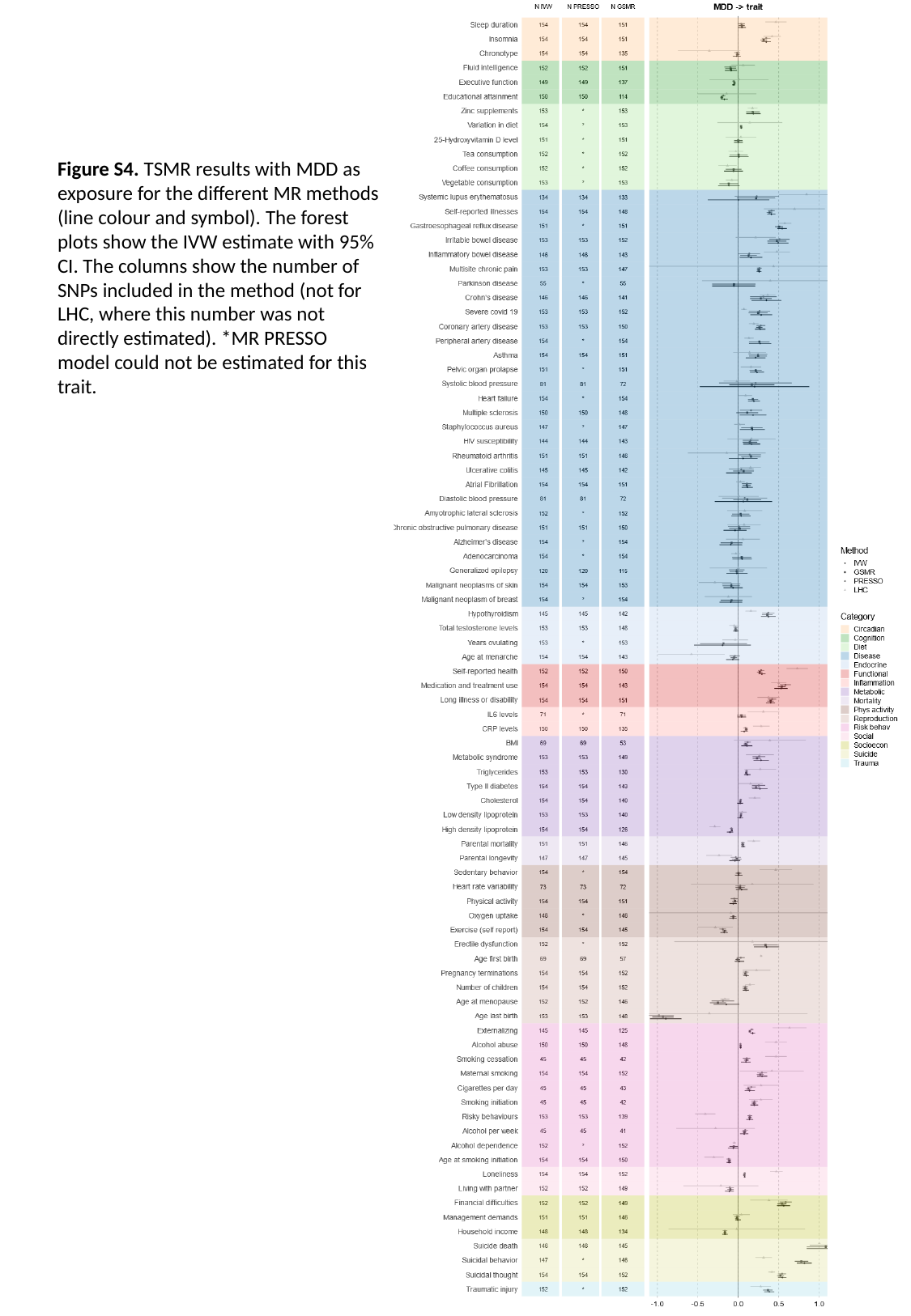

Figure S4. TSMR results with MDD as exposure for the different MR methods (line colour and symbol). The forest plots show the IVW estimate with 95% CI. The columns show the number of SNPs included in the method (not for LHC, where this number was not directly estimated). *MR PRESSO model could not be estimated for this trait.

### Slide 5
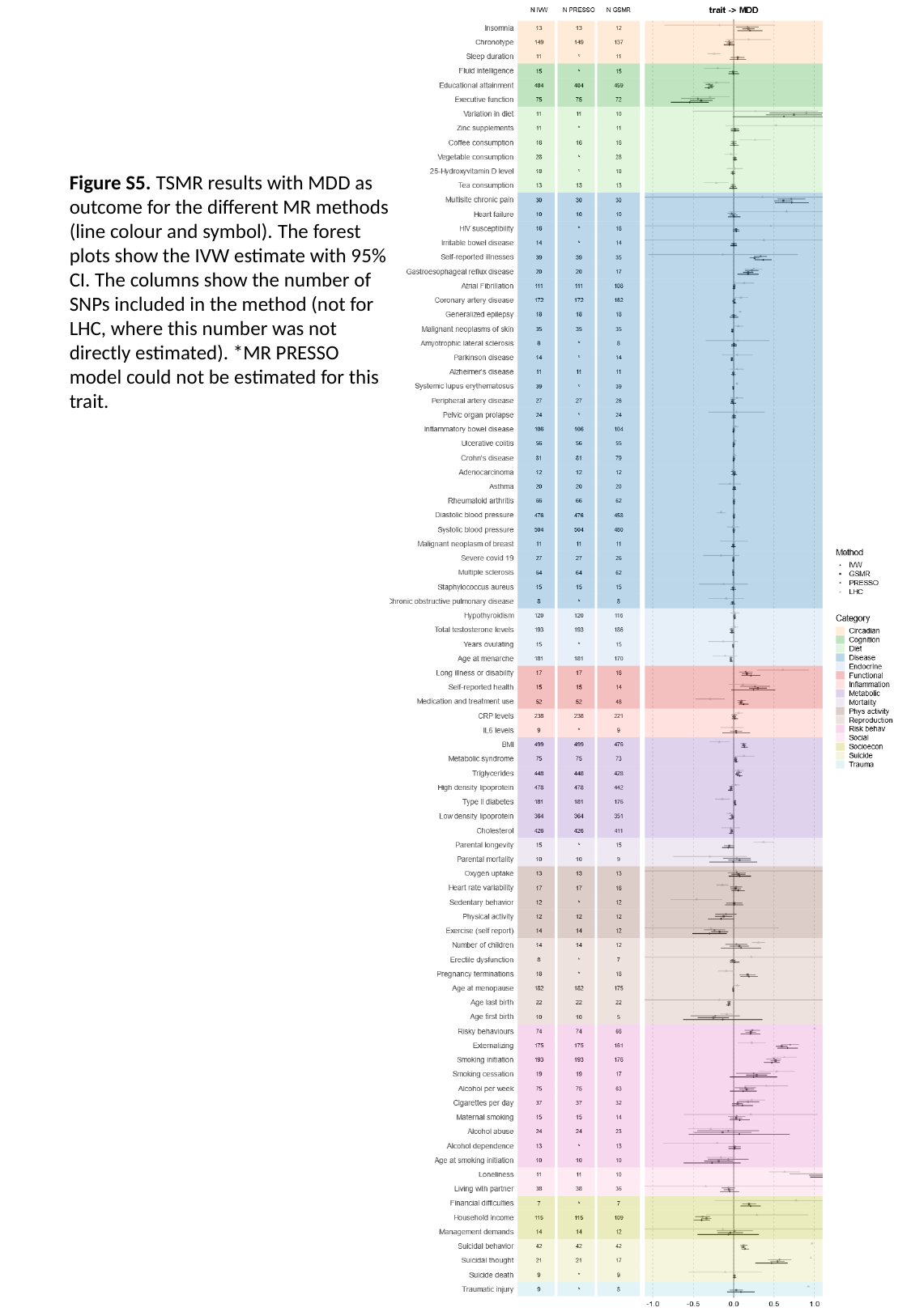

Figure S5. TSMR results with MDD as outcome for the different MR methods (line colour and symbol). The forest plots show the IVW estimate with 95% CI. The columns show the number of SNPs included in the method (not for LHC, where this number was not directly estimated). *MR PRESSO model could not be estimated for this trait.

### Slide 6
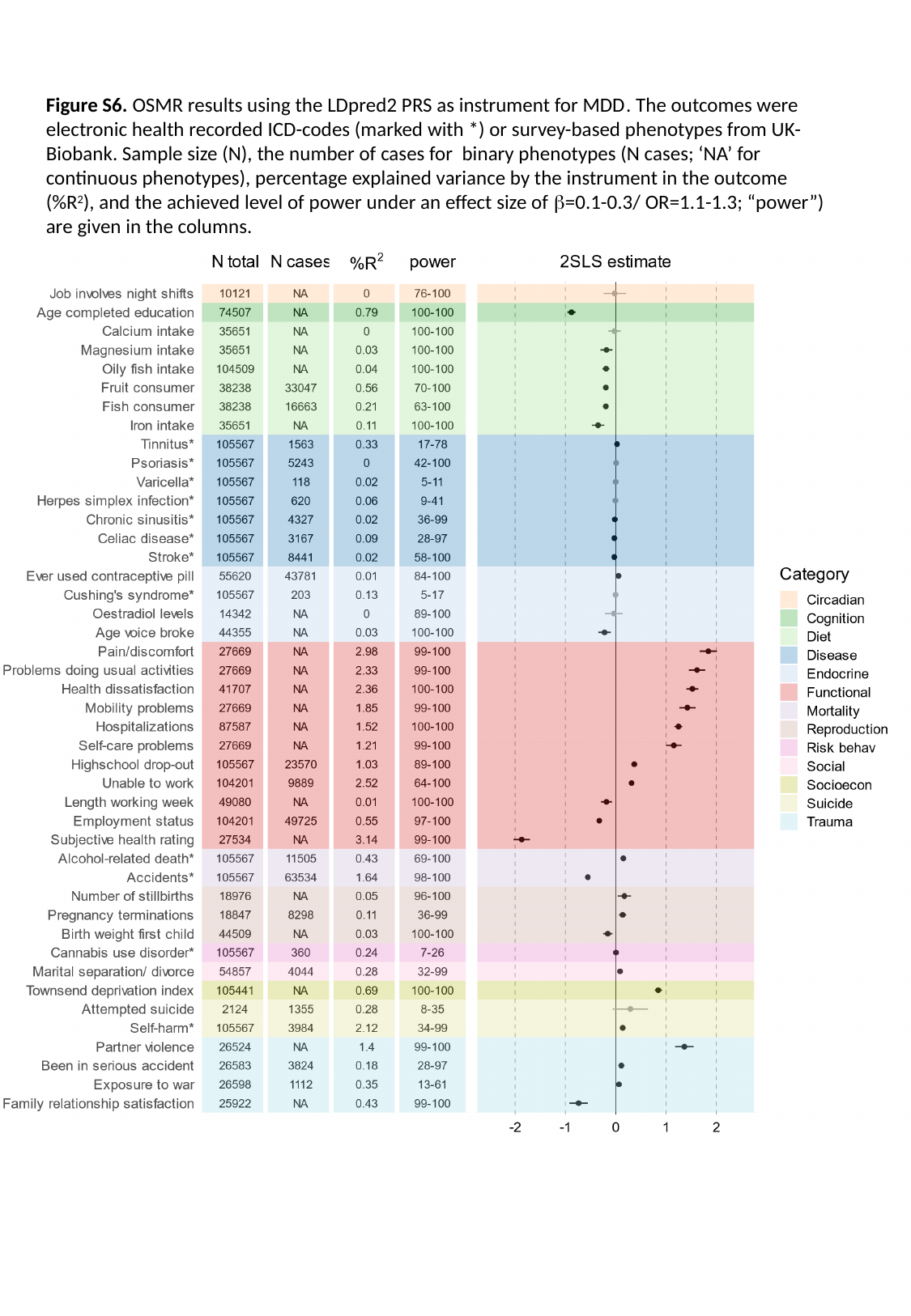

Figure S6. OSMR results using the LDpred2 PRS as instrument for MDD. The outcomes were electronic health recorded ICD-codes (marked with *) or survey-based phenotypes from UK-Biobank. Sample size (N), the number of cases for binary phenotypes (N cases; ‘NA’ for continuous phenotypes), percentage explained variance by the instrument in the outcome (%R^2^), and the achieved level of power under an effect size of β=0.1-0.3/ OR=1.1-1.3; “power”) are given in the columns.
