## Supplementary information for "Causes and consequences of major depressive disorder: An encompassing Mendelian randomization study"

1. **Methods for GWAS trait selection**

GWAS catalog^1^, GWAS ATLAS^2^, the fastGWA repository^3^, and PubMed were searched for GWAS on the identified traits by the literature review. Psychiatric traits were not included as risk factor or outcome, given the likelihood of comorbidity and pleiotropic associations with MDD. The exception is suicide, which is considered a key outcome in MDD. We manually categorized the identified traits into 18 categories, that are listed alongside their source in Table S2-S3 and are used throughout the results section to aid interpretation.

We selected GWAS for analysis if summary statistics were publicly available and N≥10,000 for continuous traits, or N cases≥5,000 for binary traits, as these sample sizes are generally needed to yield sufficient power in MR analyses^4^. Next, the summary statistics were analyzed with LDscore regression to determine SNP-based heritability. Traits that had a SNP-based heritability that was not significant and/or fell below 3% were excluded. Traits with at least 5 GW-SNPs as reported in the source GWAS were selected. The N, heritability, and number of instruments criteria serve to ensure power for the MR analyses and are based on observations and recommendations from seminal MR papers, e.g. ^5–7^. In addition, we focused on European ancestry GWAS because of the limited availability of other-ancestry GWAS. If there were only trans ancestry summary statistics available for a trait, they were still included if the majority of the sample was European. Because of differing allele frequencies in different populations, results relying on mixed ancestry GWAS need to be interpreted with caution (Table S2). Because most of the considered traits are binary, effect sizes are comparable in the exposure and outcome GWASs. Wherever this is not the case (or wherever there are substantial differences in the operationalization of the binary traits), GWAS betas cannot be interpreted in the same manner. Thus, MR estimates are not directly compared between trait pairs.

2. **Methods for Two-Sample MR follow-up analyses**

Causal interpretation of MR findings is supported if the analyses meet 3 core assumptions, including relevance (the genetic instruments reliably capture the exposure), independence (the genetic instruments are not related to the outcome through an unmeasured confounder), and exclusion restriction (the instruments are only related to the outcome through their association with the exposure). The first assumption is met through selecting variants with a strong association in GWAS with sufficient power (based on SNP-heritability and sample size). A wide range of sensitivity analyses is employed to fulfill the other assumptions, mostly boiling down to various ways of dealing with pleiotropy. These methods combined should account for those sources of pleiotropy that can be measured, although it will remain uncertain to what extent pleiotropy stemming from other, unmeasurable sources play a role (e.g., assortative mating or dynastic effects).

As a first sensitivity check, we performed Steiger filtering of the standard IVW estimate and excluded instrument SNPs that explained more variance in the outcome than in the exposure at *p*<.05 to counter horizontal pleiotropic effects. Second, to check for pleiotropy, we estimated the MR Egger intercept, and flagged results that were significantly pleiotropic according to this parameter. The Q-heterogeneity statistic was reported to indicate if the effect was uniform across the instrument SNPs, or driven by only some of them (which can also be an indication of pleiotropy). To assess the threat of weak instrument bias we reported the *F*-statistic and excluded instruments with an *F*<10. We also reported weighted median and mode results, that are more robust to weak or pleiotropic instruments.

As an alternative TSMR method, we re-estimated the effects using Generalised Summary-data-based Mendelian Randomisation (GSMR) as implemented in the Genome-wide Complex Trait Analysis (GCTA) software package^8^. GSMR is a powerful adaptation of the TSMR method that models the linkage disequilibrium structure between SNPs to allow for correlated instrumental SNPs (avoiding the power loss associated with excluding many instruments). We applied HEIDI outlier filtering in the GSMR analysis, which removes pleiotropic instruments in a manner equivalent to Steiger filtering. Furthermore, we repeated the analyses in Latent Heritable Confounder MR (LHC-MR)^9^. This method aims to adjust for unmeasured confounders by estimating latent heritable factors. Crucial for our purposes, it is robust to sample overlap, which was unavoidable in this study due to the wide range of selected traits. As a trade-off, the LHC-MR effect estimates seem to be less precise (i.e., have wider confidence intervals). Finally, we performed MR-PRESSO analyses using the MRPRESSO R-package. MR-PRESSO is a tool to correct for pleiotropic outliers relying on leave-one-out analysis, thereby retaining more statistical power than MR-Egger^10.^

3. **STROBE-MR checklist of recommended items to address in reports of Mendelian randomization studies**^1^ ^2^

| **Item No.** | **Section** | **Checklist item** | **Page No.** | **Relevant text from manuscript** |
| --- | --- | --- | --- | --- |
| 1 | **TITLE and ABSTRACT** | Indicate Mendelian randomization (MR) as the study’s design in the title and/or the abstract if that is a main purpose of the study | 1 | Title, methods section abstract |
|  | **INTRODUCTION** |  |  |  |
| 2 | **Background** | Explain the scientific background and rationale for the reported study. What is the exposure? Is a potential causal relationship between exposure and outcome plausible? Justify why MR is a helpful method to address the study question | 2, 6, Table S1 | Brief overview of risk factors and outcomes in introduction; comprehensive literature review in results section |
| 3 | **Objectives** | State specific objectives clearly, including pre-specified causal hypotheses (if any). State that MR is a method that, under specific assumptions, intends to estimate causal effects | 2 | Last paragraph introduction |
|  | **METHODS** |  |  |  |
| 4 | **Study design and data sources** | Present key elements of the study design early in the article. Consider including a table listing sources of data for all phases of the study. For each data source contributing to the analysis, describe the following: |  | *Due to the scale of this study, it was not feasible to offer a high level of detail for all aspects listed here. Key study features have been reported in Table S2.* |
|  | a) | Setting: Describe the study design and the underlying population, if possible. Describe the setting, locations, and relevant dates, including periods of recruitment, exposure, follow-up, and data collection, when available. | NA | NA |
|  | b) | Participants: Give the eligibility criteria, and the sources and methods of selection of participants. Report the sample size, and whether any power or sample size calculations were carried out prior to the main analysis | 3 | Studies had to meet strict inclusion criteria for instrument selection to ensure power |
|  | c) | Describe measurement, quality control and selection of genetic variants | 3-4 | Automated selection and QC procedures were applied as implemented in the TSMR package |
|  | d) | For each exposure, outcome, and other relevant variables, describe methods of assessment and diagnostic criteria for diseases | NA | NA |
|  | e) | Provide details of ethics committee approval and participant informed consent, if relevant | NA | Studies have all been conducted under relevant ethical approval. |
| 5 | **Assumptions** | Explicitly state the three core IV assumptions for the main analysis (relevance, independence and exclusion restriction) as well assumptions for any additional or sensitivity analysis | Sup Info 2 | Assumptions and sensitivity analyses conducted to address them have been described |
| 6 | **Statistical methods: main analysis** | Describe statistical methods and statistics used |  |  |
|  | a) | Describe how quantitative variables were handled in the analyses (i.e., scale, units, model) | NA | Due to the scale of the study, this has not been described for all traits, though care was taken to not directly compare effect sizes between traits (Sup Info 1) |
|  | b) | Describe how genetic variants were handled in the analyses and, if applicable, how their weights were selected | 3-4 | Described under TSMR methods |
|  | c) | Describe the MR estimator (e.g. two-stage least squares, Wald ratio) and related statistics. Detail the included covariates and, in case of two-sample MR, whether the same covariate set was used for adjustment in the two samples | 4 | For TSMR, the covariate set in both samples has not been detailed, though almost all GWASs have controlled for age/ birth year, sex, and ancestry |
|  | d) | Explain how missing data were addressed | Sup Info 3.1 | Only relevant for OSMR analysis |
|  | e) | If applicable, indicate how multiple testing was addressed | 3,5 | Bonferroni for LDSC, FDR for MR analyses |
| 7 | **Assessment of assumptions** | Describe any methods or prior knowledge used to assess the assumptions or justify their validity | Sup Info 2 |  |
| 8 | **Sensitivity analyses and additional analyses** | Describe any sensitivity analyses or additional analyses performed (e.g. comparison of effect estimates from different approaches, independent replication, bias analytic techniques, validation of instruments, simulations) | Sup Info 2 |  |
| 9 | **Software and pre-registration** |  |  |  |
|  | a) | Name statistical software and package(s), including version and settings used | Sup Info 2 |  |
|  | b) | State whether the study protocol and details were pre-registered (as well as when and where) | 3 | Under ‘Trait selection’ in Methods section |
|  | **RESULTS** |  |  |  |
| 10 | **Descriptive data** |  |  |  |
|  | a) | Report the numbers of individuals at each stage of included studies and reasons for exclusion. Consider use of a flow diagram | Table S2, S6-7 |  |
|  | b) | Report summary statistics for phenotypic exposure(s), outcome(s), and other relevant variables (e.g. means, SDs, proportions) | Table S2 | Due to the scope of the study, we limit ourselves to reporting SNP-based heritability per GWAS |
|  | c) | If the data sources include meta-analyses of previous studies, provide the assessments of heterogeneity across these studies | NA | Heterogeneity reports can be found in the source GWASs, referenced in Table S2 |
|  | d) | For two-sample MR:  i.  Provide justification of the similarity of the genetic variant-exposure associations between the exposure and outcome samples  ii.  Provide information on the number of individuals who overlap between the exposure and outcome studies | Sup Info 1-2 | i. Explained in Sup Info 1  ii. To limit sample overlap, we excluded UK-Biobank. In addition, we performed LHC-MR which can deal with sample overlap (Sup Info 2) |
| 11 | **Main results** |  |  |  |
|  | a) | Report the associations between genetic variant and exposure, and between genetic variant and outcome, preferably on an interpretable scale | NA | Source GWAS are referenced in Sup Table S2 |
|  | b) | Report MR estimates of the relationship between exposure and outcome, and the measures of uncertainty from the MR analysis, on an interpretable scale, such as odds ratio or relative risk per SD difference | Table S5-6 | Because of the range of considered traits, we have not converted estimates to interpretable scales. Estimates are accompanied by standard errors and a range of sensitivity estimates and robustness parameters instead |
|  | c) | If relevant, consider translating estimates of relative risk into absolute risk for a meaningful time period | NA | NA |
|  | d) | Consider plots to visualize results (e.g. forest plot, scatterplot of associations between genetic variants and outcome versus between genetic variants and exposure) | Fig 3-4 |  |
| 12 | **Assessment of assumptions** |  |  |  |
|  | a) | Report the assessment of the validity of the assumptions | 8,9,11, Table S5-6, Fig 3-4 | Reports of sensitivity and robustness checks aimed at checking assumptions are integrated in results text, figures, and tables |
|  | b) | Report any additional statistics (e.g., assessments of heterogeneity across genetic variants, such as *I^2^*, Q statistic or E-value) | Fig 3-4, Table S5-6 | Power estimates, Q, I^2^, etc. are reported in results figures and tables |
| 13 | **Sensitivity analyses and additional analyses** |  |  |  |
|  | a) | Report any sensitivity analyses to assess the robustness of the main results to violations of the assumptions | 8,9,11, Table S5-6, Fig 3-4, Sup figures | Sensitivity analyses are integrated in results text, figures, and tables |
|  | b) | Report results from other sensitivity analyses or additional analyses | 8,9,11, Table S5-6, Fig 3-4, Sup figures | Sensitivity analyses are integrated in results text, figures, and tables |
|  | c) | Report any assessment of direction of causal relationship (e.g., bidirectional MR) | 8,9, Table S5, Fig 3 | TSMR analyses have been conducted bidirectionally |
|  | d) | When relevant, report and compare with estimates from non-MR analyses | Fig 4 | Observational regression analyses are reported for OSMR |
|  | e) | Consider additional plots to visualize results (e.g., leave-one-out analyses) | Sup figures | Broader overview of sensitivity analyses are presented in Fig S1-6 |
|  | **DISCUSSION** |  |  |  |
| 14 | **Key results** | Summarize key results with reference to study objectives | 13 |  |
| 15 | **Limitations** | Discuss limitations of the study, taking into account the validity of the IV assumptions, other sources of potential bias, and imprecision. Discuss both direction and magnitude of any potential bias and any efforts to address them | 14 |  |
| 16 | **Interpretation** |  |  |  |
|  | a) | Meaning: Give a cautious overall interpretation of results in the context of their limitations and in comparison with other studies | 13-14 |  |
|  | b) | Mechanism: Discuss underlying biological mechanisms that could drive a potential causal relationship between the investigated exposure and the outcome, and whether the gene-environment equivalence assumption is reasonable. Use causal language carefully, clarifying that IV estimates may provide causal effects only under certain assumptions | 13-14 |  |
|  | c) | Clinical relevance: Discuss whether the results have clinical or public policy relevance, and to what extent they inform effect sizes of possible interventions | 14 |  |
| 17 | **Generalizability** | Discuss the generalizability of the study results (a) to other populations, (b) across other exposure periods/timings, and (c) across other levels of exposure | 14 |  |
|  | **OTHER INFORMATION** |  |  |  |
| 18 | **Funding** | Describe sources of funding and the role of funders in the present study and, if applicable, sources of funding for the databases and original study or studies on which the present study is based | 15 |  |
| 19 | **Data and data sharing** | Provide the data used to perform all analyses or report where and how the data can be accessed, and reference these sources in the article. Provide the statistical code needed to reproduce the results in the article, or report whether the code is publicly accessible and if so, where | 15 |  |
| 20 | **Conflicts of Interest** | All authors should declare all potential conflicts of interest |  |  |

This checklist is copyrighted by the Equator Network under the Creative Commons Attribution 3.0 Unported (CC BY 3.0) license.

1.	Skrivankova VW, Richmond RC, Woolf BAR, Yarmolinsky J, Davies NM, Swanson SA, et al. Strengthening the Reporting of Observational Studies in Epidemiology using Mendelian Randomization (STROBE-MR) Statement. JAMA. 2021;under review.

2.	Skrivankova VW, Richmond RC, Woolf BAR, Davies NM, Swanson SA, VanderWeele TJ, et al. Strengthening the Reporting of Observational Studies in Epidemiology using Mendelian Randomisation (STROBE-MR): Explanation and Elaboration. BMJ. 2021;375:n2233.

4. **Methods for One-Sample MR analyses**
   4.1. **Outcome variable definitions**

For the OSMR analyses, we selected traits that were not captured with GWAS but were measured in the UK-Biobank. For disease traits (including MDD) we relied on hospital inpatient records (datafield 41270), that were available in maximum N=10,121 individuals for whom MDD case status could be derived. All diagnoses were coded according to International Statistical Classification of Diseases 10^th^ edition. The codes used to define outcomes are summarized in Table S3. For all diagnoses, we looked at lifetime prevalence, because prevalence rates were too low to distinguish between a diagnosis before or after MDD onset while retaining statistical power, and because the self-report traits were also measured without taking into account MDD onset. MDD was defined as having a lifetime diagnosis coded F32 (depressive episode) or F33 (recurrent depression). All individuals without the target diagnosis are used as controls. The advantage of relying on electronic health records (EHR) for the disease outcomes is that we could use a similar, real-world measures to capture a range of outcomes, without having to rely on self-report and potentially arbitrary thresholds for diagnosing any condition. The potential drawback is that not all cases may have an EHR record, and some may end up in the control group. Still, given the seriousness of the considered outcomes, most cases should still be captured in EHR.

The self-report traits were cleaned following the UK-Biobank coding scheme. Continuous variables were winsorized, such that values more than 4 standard deviations from the average were set to the maximum (for positive outliers) or minimum value (for negative outliers), so that the outliers could not distort the results, but were not ignored either. The continuous variables were furthermore standardized to ensure comparability across outcomes. Ordinal (non-binary) outcomes were treated as continuous variables in the analysis.

4.2. **Sensitivity analysis with LDpred2 PRS**

Because our PRS instrument suffered from weak instrument bias, we repeated the analyses using a PRS created with LDpred2. LDpred2 by default includes information from many more SNPs, making this instrument more likely to be pleiotropic, even if it is more powerful. We preformed LDpred2 on automatic mode, using the SNPs from the European ancestry HapMap3 reference panel. Missing genotypes were imputed using the mean value from the reference data. The prior for the proportion of causal variants that were predicted to contribute to the phenotype was set at 0.2, which is the value suggested by the LDpred2 authors. Repeating procedures for the classic PRS, the LDpred2 PRS was standardized, and sex, birth year (log transformed), and the standardized first 10 principal components for genetic ancestry were regressed out.

4.3. **Instrument strength**

Next, we estimated R^2^ (variance explained in observed MDD by the instrument PRS) and *F*, which reflects instrument strength^4^. For *F*, we used a formula taking into account the sample size *N* and the number of instrument SNPs *k*:

$$F=\frac{R^{2}(N-1-k)}{\left( 1-R^{2} \right)k}$$

Our main results relied on the standard Plink PRS, for which *k* was the number of independent instrument SNPs for MDD as used in TSMR (*k=*150). For the sensitivity analyses using the LDpred2 PRS, the number of independent instrument SNPs *k* cannot be derived, since LDpred2 does not use clumping or *p*-thresholding in the traditional sense. To approach *k,* we counted the clumped genome-wide significant hits at *p*<5E-8, R^2^=0.01, and distance <1,000 kb using the clumping function from the TwoSampleMR package, relying on European ancestry reference data from 1000 Genomes. This resulted in *k=*203.

4.4. **Methods for observational OLS regression analysis**

To facilitate comparison of the 2SLS results with observational associations, we performed regression analyses on the observed data, again regressing each outcome on MDD, while controlling for sex and birth year. For continuous outcomes, we used linear regression. For binary outcomes, we used logistic regression.

4.5. **Per-SNP association tests for sensitivity follow-up analyses**

To be able to use TSMR methods for the sensitivity analyses (including IVW, MR Egger, and weighted median and mode) association analyses needed to be performed between each SNP in the PRS and each outcome. For these analyses we used Plink --linear for continuous outcomes and --logistic for binary outcomes while controlling for sex, birth year, and the first 10 principal components capturing population stratification (as provided by UK-Biobank). Because the number of included SNPs was limited and the number of estimated parameters was high, association staetistics could not be derived for 4 (out of 48) traits. For the remainder, sensitivity analyses could be performed relying on the same methods as used for TSMR, although the number of SNPs was insufficient to estimate the weighted median for 22 outcomes.

5. **Results for One-Sample MR using the LDpred2 PRS instrument**

We repeated the OSMR using a putatively more powerful SNP instrument created using LDpred2. Overall, results were similar to that from the classic PRS, but confidence intervals were more narrow. The strongest negative effects were observed for age at completion of education, subjective health rating, accidents, and the strongest positive effects for pain, problems with usual activities, and health dissatisfaction. Interestingly, some of the small effects that were not significant in the OLS were significant in the OSMR using the LDPred2-based instrument (i.e., magnesium intake, use of the contraceptive pill, pregnancy terminations, birth weight of first child, war exposure, and accidents), suggesting that there may be small causal effects that OLS cannot detect due to confounding. The PRS explained high amounts of variance in MDD (Nagelkerke’s R^2^=7.1%, which is almost the complete SNP-heritability), and there was no weak instrument bias (average *p*=3.7E-176). The Wu-Hausman again indicated that OSMR estimates were preferrable in most analyses that showed a significant association (average *p*=.06). Assuming (conservatively) that the LDpred2 PRS relied on approximately *k*=203 SNP effects, the instrument strength was estimated at F=32.0, and there was hardly evidence for pleiotropy (only celiac disease again showed a significant MR-Egger intercept). Thus, using the LDpred2-based PRS resulted in better analytical power and did not detectably increase pleiotropy. Conceptually, more pleiotropy must have been introduced due to LDpred2’s reliance on the entire genome in estimating the signal. Methodological follow-up research is warranted to establish if increasing the amount of signal captured by the PRS is a viable approach to increasing the power in OSMR analyses, while avoiding increases in pleiotropy.
